# ENDORSE: a prognostic model for endocrine therapy response in advanced estrogen-receptor positive breast cancers

**DOI:** 10.1101/2021.02.03.21251035

**Authors:** Aritro Nath, Adam L. Cohen, Andrea H. Bild

## Abstract

Endocrine therapy remains the primary treatment for advanced and metastatic estrogen receptor-positive (ER+) breast cancers. Patients who progress on endocrine therapy may benefit from add-on treatment targeting the PI3K/MTOR signaling pathways or by switching to chemotherapy. However, these options are only available after progression on first-line treatment with endocrine therapy. In the absence of reliable prognostic tests for advanced ER+ breast cancers, it is currently not possible to stratify patients into pertinent treatment arms at the baseline. To address this, we have developed a low-dimensional endocrine response signature (ENDORSE) model for advanced ER+ breast cancers. The ENDORSE model was developed using the baseline tumor transcriptomes and long-term survival outcomes of >800 invasive ER+ breast cancers and predicts the risk of death on endocrine therapy. ENDORSE was validated in multiple clinical trial datasets for endocrine therapy response in metastatic ER+ breast cancers and demonstrated superior predictive performance over clinical factors and published gene signatures. Our results show that ENDORSE is a reproducible and accurate prognostic model for endocrine therapy response in advanced and metastatic ER+ breast cancers.

## INTRODUCTION

Breast cancer is the most common form of cancer globally, with more than two million cases diagnosed in 2020^1^. Pathogenesis and classification of breast cancer is based on the presence or absence of estrogen receptor alpha (ER), progesterone receptor (PR) and human growth factor-neu receptor (HER2). These subtypes guide the selection of systemic therapy for breast cancer patients. More than 70% of breast cancers express ER and are negative for HER2 (ER+/HER2-)^2, 3^. The primary systemic therapy for ER+/HER2- breast cancer is endocrine therapy, which counters the growth of tumors by targeting their dependency on estrogen signaling^4^. This includes selective estrogen receptor modulators (SERMs) such as tamoxifen and selective estrogen receptor degraders (SERDs) such as fulvestrant that directly prevent ER activation, or aromatase inhibitors like exemestane and anastrozole that reduce circulating levels of estrogen in the body ^5, 6^. Endocrine therapy substantially reduces the risk of recurrence within 5- years, although chemotherapy may be recommended for some patients with high risk of recurrence. While clinicopathological features are not reliable predictors of recurrence risk, gene expression-based genomic tests that predict the risk of recurrence can aid in deciding whether the benefit of adding chemotherapy outweighs its side effects in certain patients^7, 8^. These biomarkers are have been validated and recommended for clinical use only in early stage, node-negative cancers based on guidelines from the American Society of Clinical Onoclogy and European Group on Tumor Markers ^9, 10^.

Locally advanced and metastatic ER+ breast cancers often develop resistance to endocrine therapy with significantly higher rates of recurrence and death compared to early-stage disease. Despite these challenges, single-agent endocrine therapy or in combination with CDK4/6 inhibitors remains the primary systemic therapy recommended for locally advanced and metastatic breast cancers^11^. Patients may benefit from the addition of targeted inhibitor against the mTOR or PI3K pathways^12, 13^ or switching to chemotherapy^11^. However, these treatment options are recommended for consideration only upon progression on endocrine therapy, according to the American Society for Clinical Oncology ^14^, National Comprehensive Cancer Network ^15, 16^ and European Society for Medical Oncology ^17^ clinical practice guidelines. Therefore, the ability to predict the potential benefit from first-line endocrine therapy may be crucial for locally advanced and metastatic ER+ breast cancers that may benefit from continued endocrine therapy, a combination treatment or chemotherapy as the primary treatment strategy.

Unlike early stage, node-negative disease, genomic tests for endocrine therapy response are not available for advanced and metastatic ER+ breast cancers. To address this limitation, a few attempts have been made so far to develop a genomic signature of endocrine response in ER+ metastatic breast cancers (ER+ MBC)^18, 19^. The TransCONFIRM trial evaluated the transcriptomes of 112 ER+/HER2-MBCs and identified a set of 37 genes that were associated with progression-free survival (PFS) of patients receiving fulvestrant^19^. Another study analyzed the transcriptomes of 140 ER+/HER2-MBC on endocrine therapy to develop SET ER/PR, an 18-gene predictive score for endocrine therapy sensitivity ^18^. While both the TransCONFIRM and SET ER/PR biomarkers predicted endocrine response in their respective training datasets, neither study performed systematic validation of their predictive signatures to demonstrate the reproducibility and accuracy in independent clinical datasets. This issue highlights a critical flaw in biomarker development pipelines and is one important reason why genomic biomarkers are infrequently translated into clinical practice^20^. Another pervasive issue hindering clinical translation arises from the reliance on a large number of predictive features in complex models that are difficult to interpret and often perform poorly in independent validation due to overfitting^21, 22^.

Here we developed ENDORSE: a low-dimensional expression-based prognostic model for endocrine therapy and systemically tested its performance and predictive ability in multiple-independent clinical trials against other diagnostic models and genomic signatures. ENDORSE was developed and trained using the tumor transcriptomes and overall survival (OS) of more than 800 ER+ breast cancers on endocrine therapy^23, 24^. We validated the ENDORSE model in multiple independent clinical trial datasets, including the TransCONFIRM and SET ER/PR trials for endocrine therapy in metastatic ER+ breast cancer. Our results show that ENDORSE reproducibly predicts endocrine response in independent validation clinical studies, and consistently outperforms all other models of endocrine therapy response in metastatic ER+ breast cancers, clinical factors, and proliferation signatures.

## RESULTS

### Developing a low-dimensional prognostic model for endocrine therapy

We developed a two-component prognostic model for endocrine therapy response using the tumor transcriptomes and long-term survival outcomes of 833 ER+/HER2- tumors that received endocrine therapy^23, 24^ (Table 1, Figure 1a). About 2 in 5 tumors in this training cohort were node-positive, while more than a third of the tumors were poorly differentiated, grade 3 tumors (Table 1). The two components included an empirical gene signature modeled on OS (median = 10 years) and a curated gene signature defining response to estrogen^25^. Figure 1a outlines the inclusion criteria for the training dataset, method for developing the empirical gene signature and the final Cox proportional hazards model based on the gene set enrichments scores (GES) of the two signatures. The empirical signature was developed by first performing a feature selection on the training dataset using a repeated cross-validation analysis of a lasso-regularized proportional hazards model. Each iteration yielded a core set of predictive features that were expanded to a correlation network. The final gene signature was derived from the consensus correlation network, defined as genes appearing in at least 50% of the cross-validation iterations (Supplementary Figure 1, Supplementary Table 1). In a bivariate Cox proportional hazards model of the training data, the empirical signature was associated with a reduction in survival probability, while the estrogen response signature was associated with improved survival (Figure 1b). The coefficients for the <u>endo</u>crine respon<u>se</u>, or ENDORSE, model was calculated using the training cohort, resulting in ENDORSE = 1.54 x (empirical signature GES) – (2.72 x estrogen response GES). The ENDORSE model could also be used to stratify the tumors based on predicted risk, for example by setting a threshold of ≥2-fold relative risk of death as “high-risk” and ≤1 risk as “low-risk”, resulting in significant differences in the Kaplan-Meier survival curves across the strata (P = 3 x 10^-14^) (Figure 1c).

**Figure 1:**
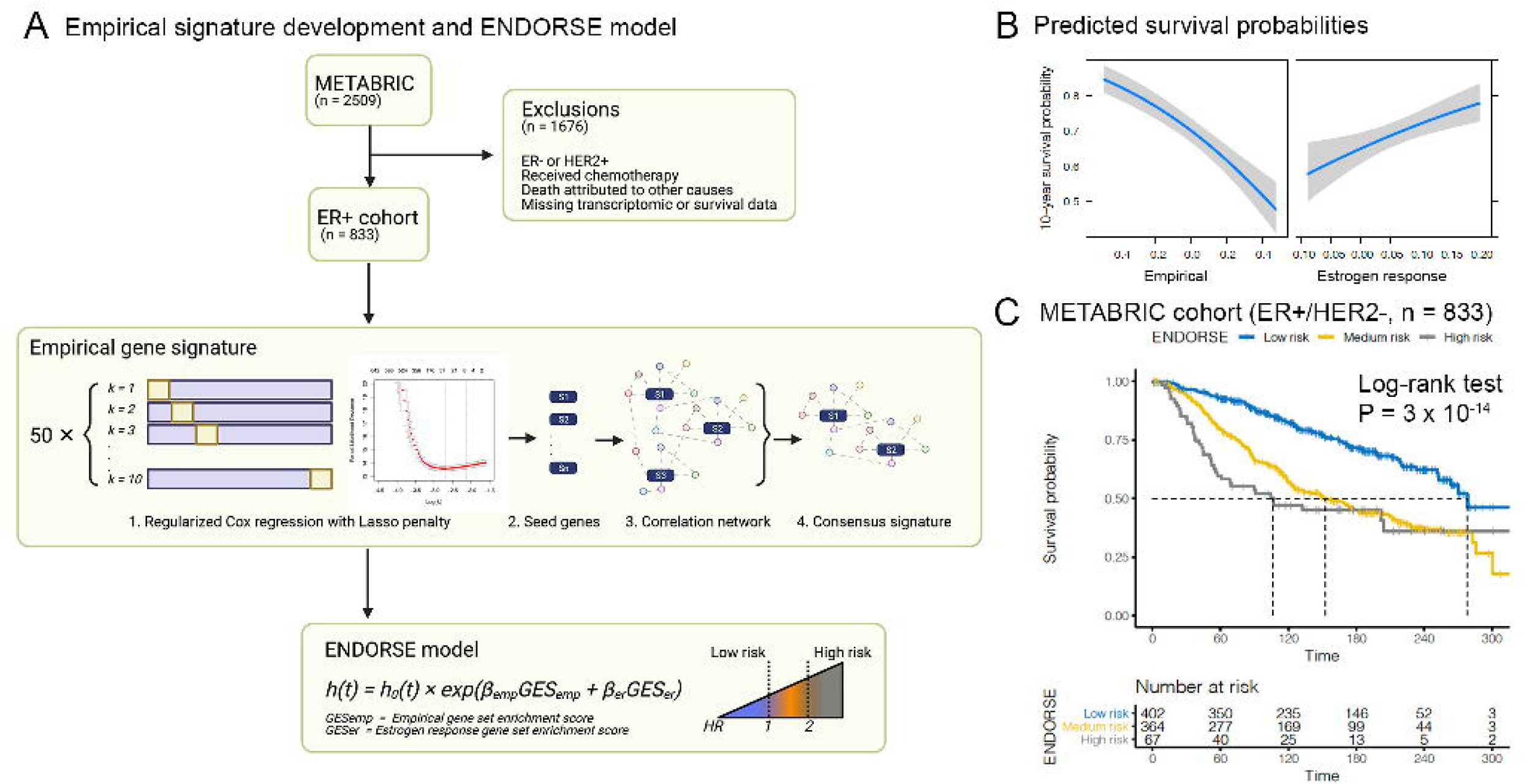
ENDORSE model development in METABRIC. A. Inclusion criteria and overall schematic of ENDORSE model development. Samples for training were selected based on ER+ status and excluded from the analysis if they were either HER2+, received chemotherapy in addition to hormone therapy, died due to other causes besides breast cancer, or were missing transcriptomic or survival data. The empirical signature was developed using a repeated cross-validation analysis framework. Each iteration of the lasso-regularized proportional hazards model generated a feature set (seed genes) predictive of OS. The seed genes were expanded to a network of intercorrelated genes, and the final empirical signature was defined by identifying a consensus set across all iterations. The two-feature ENDORSE model was then constructed using the gene set enrichment scores of the empirical signature and estrogen response signature. B. Predicted 10-year survival probabilities of the 833 ER+/HER2- METABRIC breast cancers based on a Cox proportional hazards model of gene signature enrichment scores of the empirical and estrogen response signatures as predictor variables. C. Kaplan-Meier curves and risk tables of METABRIC ER+/HER2- tumors stratified by ENDORSE. The tumors were stratified according to an ENDORSE risk score (hazard ratio) threshold of ≥2 to define high-risk, ≤1 as low risk and all other intermediate values as medium risk.

**Table 1:**
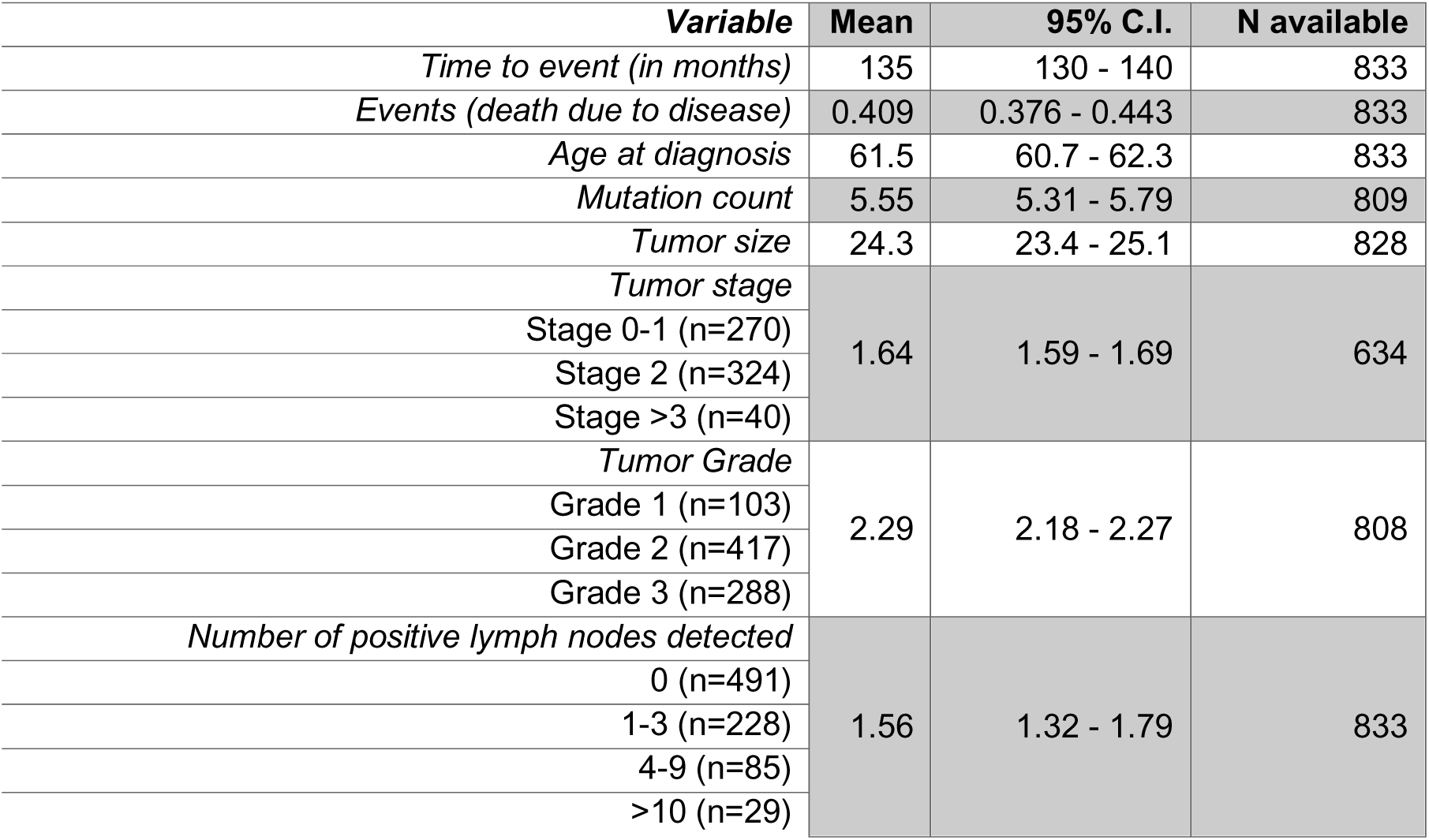
Training data patient characteristics.

### Internal performance evaluation, comparison with clinical covariates and published breast cancer signatures

We performed bootstrap resampling analyses to validate the Cox model in the training dataset (Figure 2a) and performed likelihood ratio tests (Figure 2b) to compare with other univariate prognostic models including clinical factors, proliferation index and published prognostic signatures for ER+ breast cancers. First, we compared the ENDORSE model to the univariate models based on the individual components of ENDORSE, i.e., the empirical signature and estrogen response signature. The ENDORSE model (Somer’s D or D_xy_ = 0.301) was a better fit than the empirical signature (D_xy_ = 0.296, P = 1.09 x 10^-3^) and the estrogen response signature (D_xy_ = 0.141, P = 3.93 x 10^-14^) univariate models.

**Figure 2:**
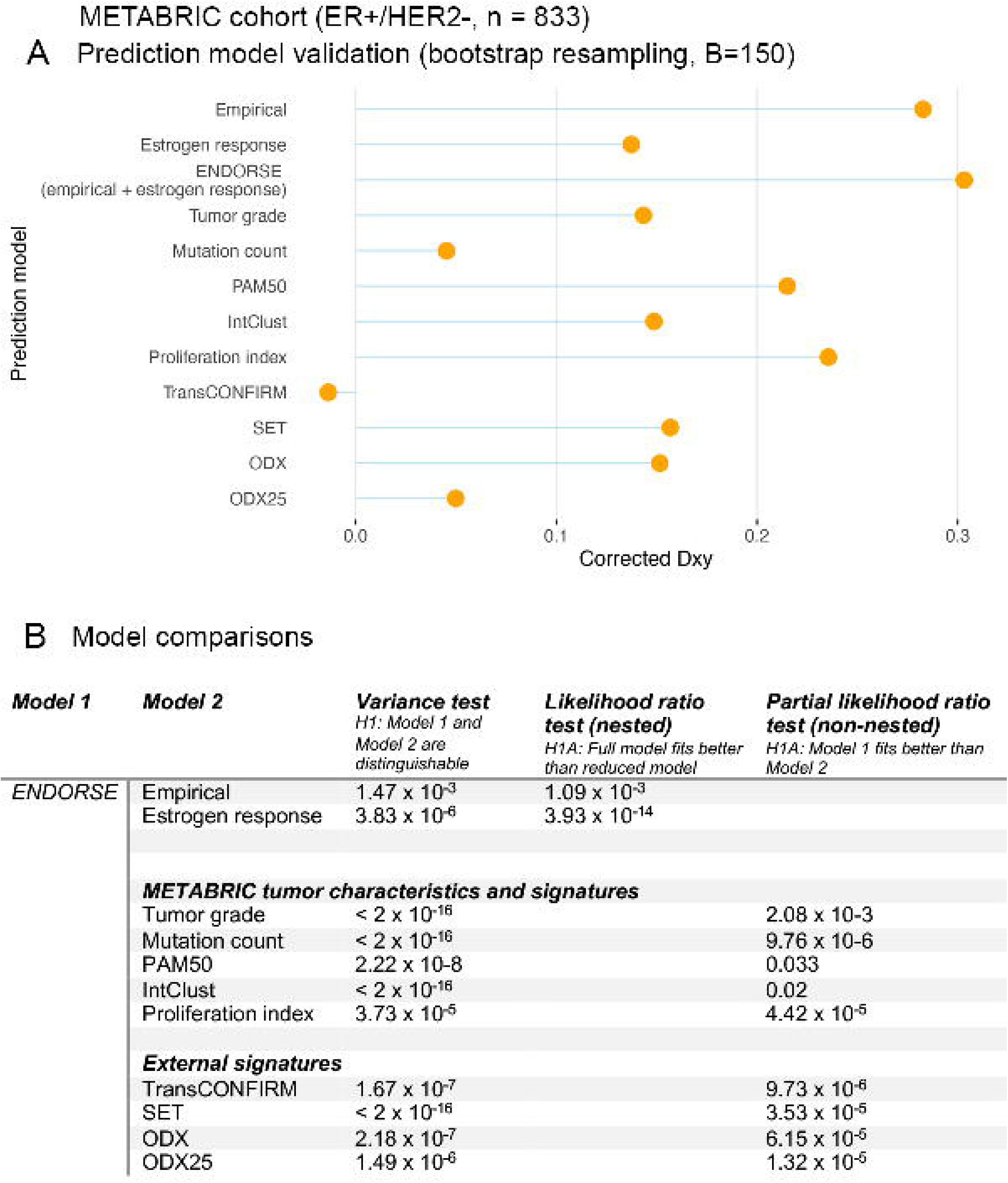
Model evaluation and comparison with other predictors. a. Lollipop plots displaying corrected Somer’s D_xy_ indices of ENDORSE and various other univariate Cox proportional hazards models. The indices were calculated using 150-fold bootstrap resampling of the training dataset. b. Table comparing the ENDORSE model with various other univariate Cox models using partial likelihood ratio tests. The comparison between the nested ENDORSE model and its two components were performed using a likelihood ratio test, while other non-nested univariate models were compared using a partial likelihood ratio test.

We then compared ENDORSE with clinical factors, such as tumor grade and mutation burden. The ENDORSE model performed better than both tumor grade (D_xy_ = 0.141, P = 2.08 x 10^-3^) and mutation count (D_xy_ = 0.059, P = 9.76 x 10^-6^). We also compared the model with a ‘meta-PCNA’ proliferation index that was reported to capture the prognostic ability of most published signatures of breast cancer^26, 27^. Again, the ENDORSE model performed significantly better than the proliferation index (D_xy_ = 0.235, P = 4.42 x 10^-5^), indicating its utility over measures of proliferation as a prognostic tool.

Next, we evaluated published prognostic signatures for breast cancers and compared their performance with ENDORSE. These signatures included PAM50, a 50-gene signature that was previously reported to be a better prognostic tool for ER+ breast cancers on endocrine therapy than clinical factors, such as histopathological classification and tumor grade^28^. A genomic classifier, IntClust, that developed by the METABRIC consortium authors and trained on the same training dataset was also included in this comparison^29^. The PAM50 model (D_xy_ = 0.220) performed better than IntClust (D_xy_ = 0.153), however the ENDORSE model outperformed both PAM50 (P = 0.033) and IntClust (P = 0.02) models.

Two previous clinical trials evaluating endocrine therapy response in metastatic ER+ breast cancers developed prognostic signatures using tumor transcriptomes. The first signature developed in the TransCONFIRM trial included 37 genes that were associated with PFS of advanced ER+ breast cancers on fulvestrant^19^. We replicated the approach described in the study by performing hierarchical clustering of the samples based on the expression levels of the 37 genes and cutting the tree to obtain two clusters. We referred to resultant clusters as the ‘TransCONFIRM’ score. The TransCONFIRM score applied to the METABRIC dataset performed poorly (D_xy_ = -.002), suggesting that the signature performed no better than a random set of genes and was unsurprisingly outperformed by ENDORSE (P = 9.73 x 10^-6^).

The second signature (SET ER/PR) was developed using tumor transcriptomes of metastatic ER+ breast cancers on endocrine therapy^18^. This signature included 18 predictive genes that were correlated with *ESR1* or *PGR* expression and normalized using 10 reference transcripts. We implemented the methods described in original study and referred to the resultant score as ‘SET’. The SET score (D_xy_ = 0.152) performed better than TransCONFIRM; however, it was also easily outperformed by ENDORSE (P = 3.53 x 10^-5^).

Finally, we calculated a surrogate based on the published formula for the 21-gene prognostic signature approved for early-stage, node-negative ER+ breast cancers ^30^. We referred to this score as ODX. We also compared a classifier that stratified samples based on 25^th^ percentile of ODX score as a proxy for the latest risk stratification threshold for this signature^8^, and referred to this score as ODX25. We found that the ODX model (D_xy_ = 0.159) was comparable to other published signatures like the SET score but the stratified ODX25 score performed poorly (D_xy_ = 0.056). Again, the ENDORSE model performed significantly better than both ODX (P = 6.15 x 10^-5^) and ODX25 (P = 1.32 x 10^-5^) models. These results show that ENDORSE is significantly better prognostic model than available gene signatures, clinical factors and proliferation index for endocrine therapy in the METABRIC dataset.

### Validation and performance evaluation in independent clinical trial datasets

To test the reproducibility and validate the performance of ENDORSE, we applied the model to the baseline transcriptomes of ER+ tumors from three independent clinical trials and compared the ENDORSE-predicted risk or strata with the outcomes reported in each trial. These independent trials also included the TransCONFIRM and SET ER/PR studies discussed earlier. So, we also compared the performance of TransCONFIRM and SET scores in their respective training datasets and also across other independent datasets.

The TransCONFIRM trial evaluated fulvestrant response in 112 advanced metastatic ER+ breast cancers previously treated with an antiestrogen^19^. While the original study developed and evaluated the performance of their 37-gene signature based on PFS, this survival data was not made available with the publication (the authors did not respond to our requests for this data). However, the study reported the post-therapy resistant or sensitive states of the tumors based on histopathological staining (Ki67 staining). Therefore, we compared the percentage of cells positive for Ki67 staining reported by in study with risk predictions from ENDORSE and other signatures (Figure 3). The percentage of cells positive of Ki67 were significantly correlated with the ENDORSE estimated risk (P = 2.5 x 10^-5^) (Figure 3a), while stratification of the patients based on the risk thresholds also showed significant difference in Ki67 staining percentage between the strata (P = 1.2 x 10-3) (Figure 3b). However, the SET score was not correlated with Ki67 staining (P = 0.3) (Figure 3c). The TransCONFIRM score that was developed on this dataset was significant (P = 0.05) but performed worse than the ENDORSE score trained on an independent dataset.

**Figure 3:**
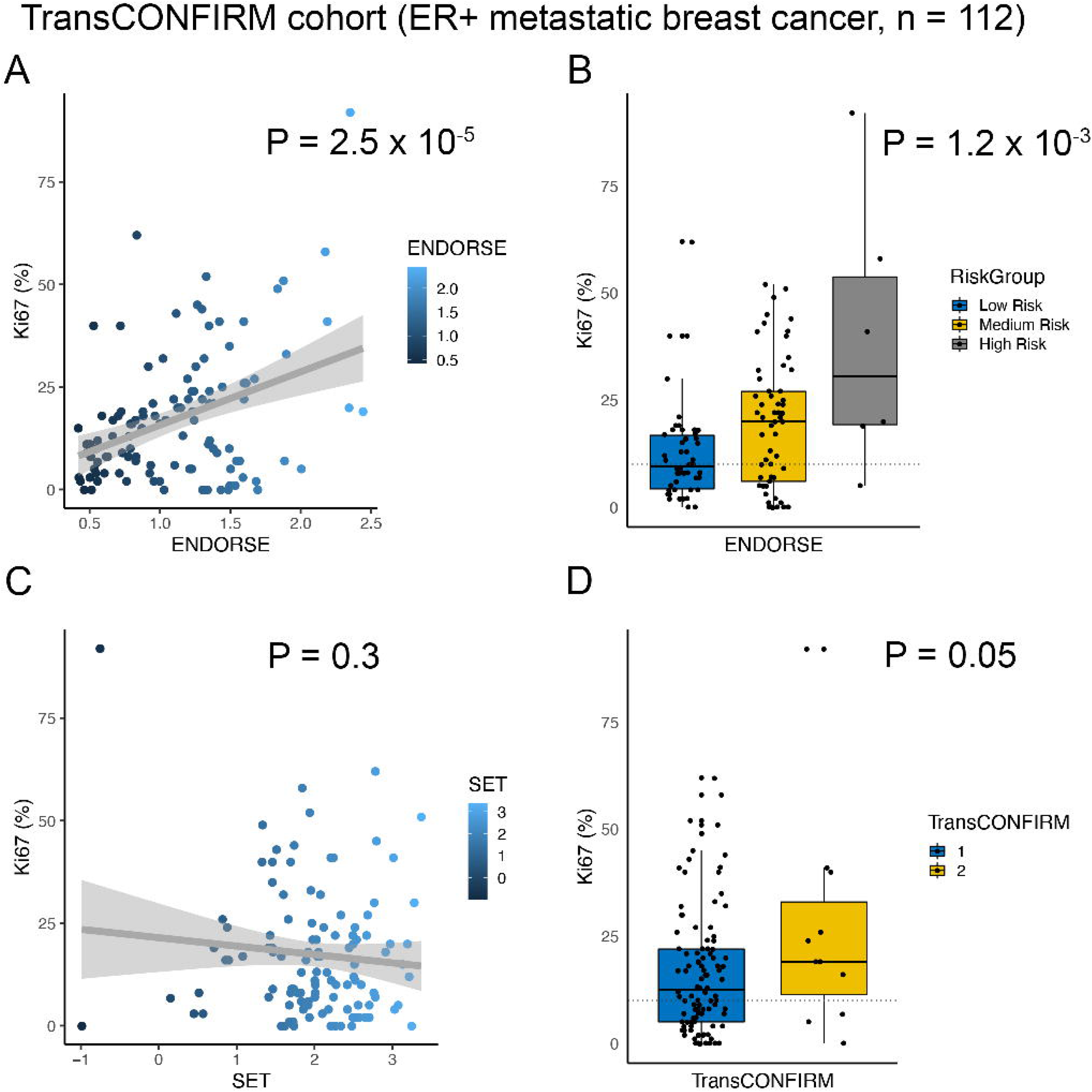
Model validation in TransCONFIRM cohort. A. Scatter plot comparing ENDORSE scores (X-axis) with trial-reported percentage of cells stained positive for Ki67 (Y-axis). Linear fit is shown as a grey line with shaded region showing 95% confidence intervals (C.I.). P-value indicates significance of the linear fit. B. Boxplot comparing Ki67 % across ENDORSE-guided patient strata. P-value indicates significance of the ANOVA model and the horizontal dotted line at 10% indicates threshold of resistance. C. Scatter plot comparing SET scores (X-axis) Ki67 % (Y-axis). Linear fit is shown as a grey line with shaded region showing 95% confidence intervals (C.I.). P-value indicates significance of the linear fit. B. Boxplot comparing Ki67 % across TransCONFIRM predicted patient strata. P-value indicates significance of the ANOVA model and the horizontal dotted line at 10% indicates threshold of resistance.

Next, we evaluated the performance of the signatures in the SET ER/PR cohort. This clinical trial reported the PFS and OS of 140 stage IV ER+ metastatic breast cancers on endocrine therapy. We compared the survival curves of the patients by stratifying them based on the ENDORSE predicted risk, median SET scores, as described in the original study, and the TransCONFIRM score. The stratification based on ENDORSE (Figure 4a) and SET (Figure 4b) scores both resulted in significant differences in the survival curves (ENDORSE P = 2 x 10^-4,^ SET P = 3 x 10^-3^). However, the TransCONFIRM score (Figure 4c) was not significant (P = 0.9). Similarly, we observed that ENDORSE (Figure 4d) and SET (Figure 4e) scores both resulted in significant differences in the PFS curves (ENDORSE P = 1 x 10^-6,^ SET P = 5 x 10^-3^), while TransCONFIRM was not significant (P = 0.2). Additionally, we compared the model fits using partial likelihood ratio tests. The SET model that was trained using the same dataset was not a better fit than the ENDORSE mode (OS P = 0.667, PFS P=0.258). The ENDORSE model was a better fit than the TransCONFIRM model in each case (OS P = 0.046, PFS P=0.038). In addition to the two metastatic ER+ breast cancer trials, we also evaluated the performance of the signatures examined data from the ACOSOG Z1031B clinical trial which evaluated neoadjuvant aromatase inhibitor (AI) treatment in Stage II or III ER+ breast cancers^31^. This study reported percentage of Ki67 staining both at the study baseline and at the end of treatment (2-4 weeks). We compared the percentage of Ki67 positive cells across cancers stratified by the ENDORSE score and found significant difference across the classes at both the baseline (P = 4.9 x 10^-9^) and at the end of treatment (P = 3 x 10^-18^) (Figure 5a). Similarly, the continuous ENDORSE scores were significantly correlated with both the baseline (P = 3.3 x 10^-15^) and end of treatment (P = 1.1 x 10^-17^) Ki67 percentage (Figure 5b). The ENDORSE scores were also significantly higher in the tumors that were classified as resistant based on clinical response (P = 4.6 x 10^-6^) (Figure 5c). In this cohort, the SET score was also significantly correlated with Ki67 percentage at the baseline (P = 2.8 x 10^-5^) and end of treatment (P = 2.2 x 10^-4^) (Figure 5d), with significant difference in the SET scores between the resistant and sensitive tumors (P = 0.05) (Figure 5e). The transCONFIRM scores were not significant at the baseline (P = 0.5) and end of treatment (Figure 5f) or between resistant and sensitive tumors (P = 0.7) (Figure 5g).

**Figure 4:**
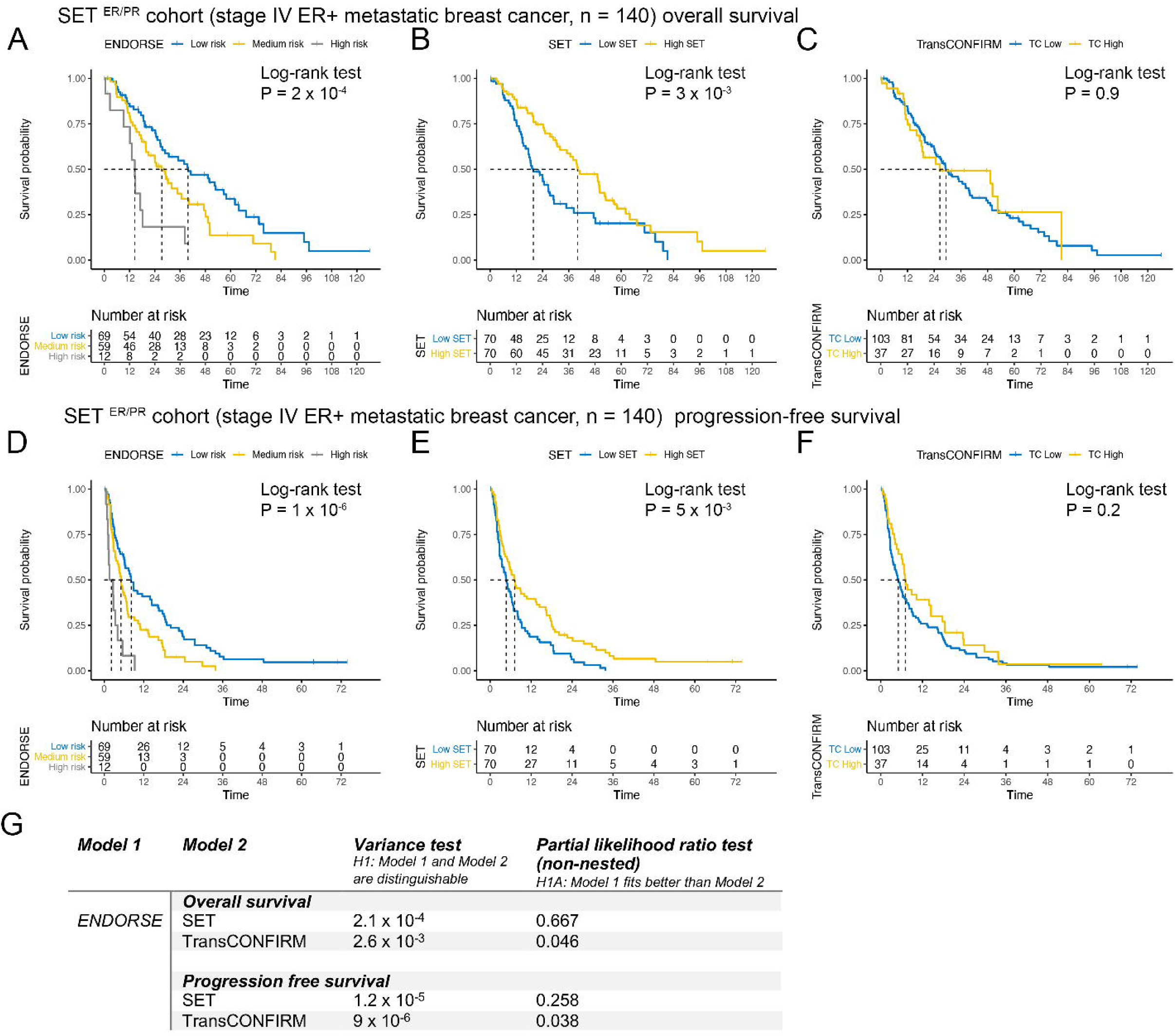
Model validation in SET ER/PR cohort. A-C. OS Kaplan-Meir curves and risk tables of SET ER/PR patients. The patients were stratified according to A. ENDORSE B. SET and C. TransCONFIRM predicted scores. P-values indicate significance of difference in survival curves based on log-rank tests. D-F PFS Kaplan-Meir curves and risk tables of SET ER/PR patients. The patients were stratified according to A. ENDORSE B. SET and C. TransCONFIRM scores. P-values indicate significance of difference in survival curves based on log-rank tests. G. Table comparing the ENDORSE overall and PFS models with SET and TransCONFIRM models using partial likelihood ratio tests for non-nested Cox models.

**Figure 5:**
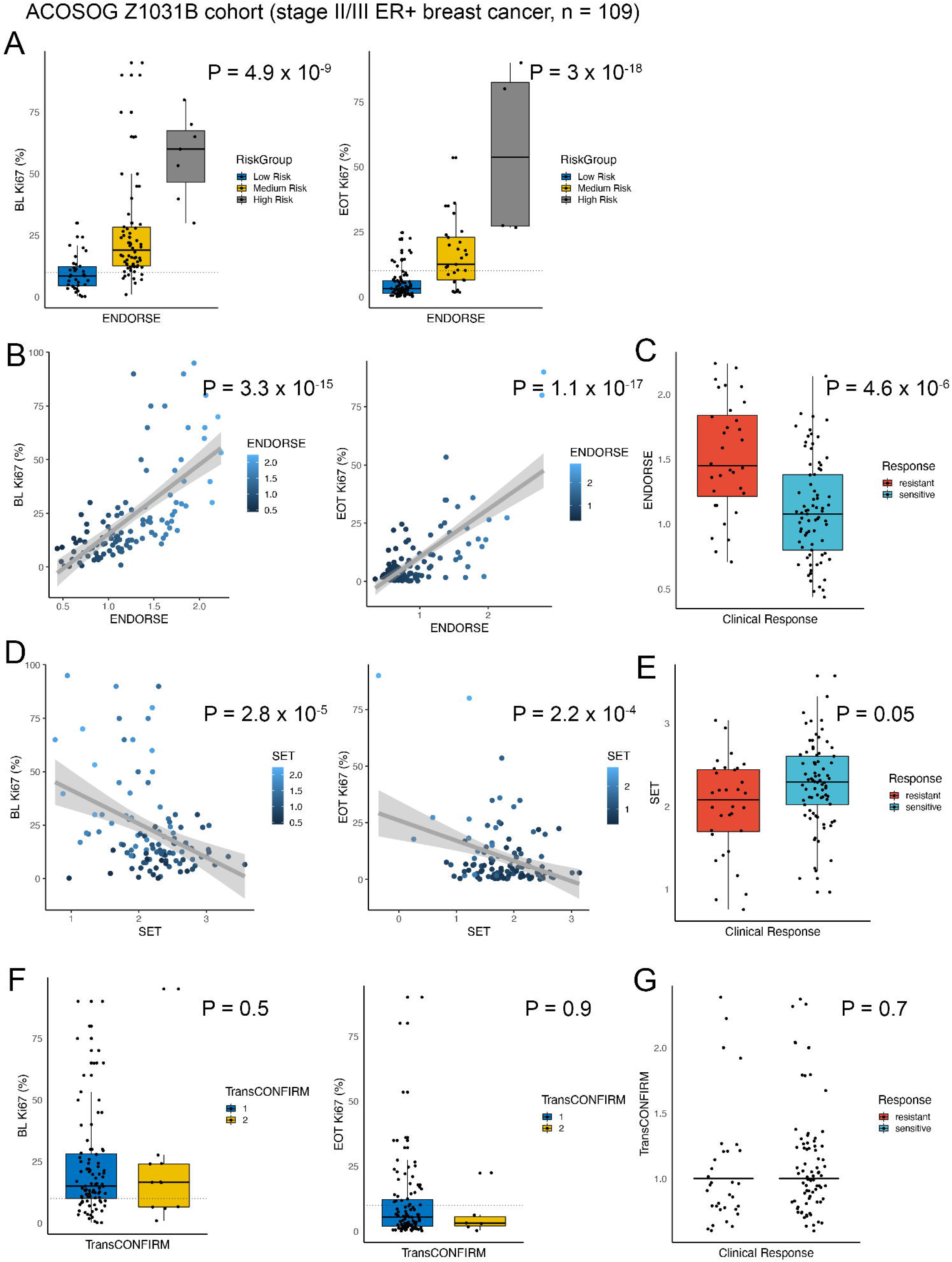
Model validation in ACOSOG Z1031B cohort. A. Boxplots comparing Ki67 % at the baseline (left panel) and end of treatment (right panel) across ENDORSE- predicted patient strata. P-value indicates significance of the ANOVA model and the horizontal dotted line at 10% indicates threshold of resistance. B. Scatter plot comparing ENDORSE scores (X-axis) and Ki67 % (Y-axis) at the baseline (left panel) and end of treatment (right panel). Linear fit is shown as a grey line with shaded region showing 95% confidence intervals (C.I.). P-value indicates significance of the linear fit. C. Boxplots comparing ENDORSE scores between patients classified as resistant or sensitive clinical response. P-value indicates significance of the ANOVA model. D. Scatter plot comparing SET scores (X-axis) and Ki67 % (Y-axis) at the baseline (left panel) and end of treatment (right panel). Linear fit is shown as a grey line with shaded region showing 95% confidence intervals (C.I.). P-value indicates significance of the linear fit. E. Boxplots comparing SET scores between patients classified as resistant or sensitive clinical response. P-value indicates significance of ANOVA model. F. Boxplots comparing Ki67 % at the baseline (left panel) and end of treatment (right panel) across TransCONFIRM-predicted patient strata. P-value indicates significance of the ANOVA model and the horizontal dotted line at 10% indicates threshold of resistance. G. Boxplots comparing TransCONFIRM predictions between patients classified as resistant or sensitive clinical response. P-value indicates significance of ANOVA model

In addition to the endocrine therapy trials in ER+ breast cancer, we also applied the ENDORSE risk estimates to stratify 429 ER-negative METABRIC breast cancers as negative controls. Kaplan-Meier analyses show no significant difference between the strata (P = 0.26, Supplementary Figure 2). This suggests that the ENDORSE model is specific to the ER+ cohort and not a general prognostic model.

### Common pathway phenotypes and somatic alterations enriched in high-risk tumors

We analyzed the pathway phenotypes enriched in each dataset to identify potential mechanisms that defined the high-risk tumors. First, we calculated the GES for 50 hallmark, 4690 curated and 189 oncogenic signatures from the METABRIC transcriptomes and fitted a generalized additive model for ENDORSE scores with each signature as the predictor (Supplementary Tables 2-4). We found multiple hallmark signatures and oncogenic pathways to be significantly associated with the ENDORSE scores (Supplementary Tables 2-4). Key enriched hallmark signatures included MTOR signaling (P = 1.03×10^-72^) and MYC targets (v2, P = 2.66×10^-83^), while key oncogenic signatures included gain in E2F1 target expression (P = 8.06×10^-302^) and loss of RB1 activity via p107 and p130 (P = 9.51×10^-137^, 1.31×10^-67^) (Supplementary Tables 2, 4). Next, we calculated the GES for the hallmark and oncogenic signatures in the three validation datasets (Supplementary Tables 5-10). We observed that pathways associated with cell-cycle progression and proliferation, along with signatures for the loss of RB-1 activity and activation of the PI3K/AKT/MTOR signaling pathways were generally enriched across the METABRIC and all the three validation datasets (Figure 6a). Similar to the training dataset, we also found gain in cell cycle progression along with MTOR signaling and E2F1 target expression to be associated with high ENDORSE scores across all datasets (Figure 6a, Supplementary Tables 5-10). The commonality of the signatures enriched across different datasets suggested similar underlying phenotypes were acquired by the high-risk tumors.

**Figure 6:**
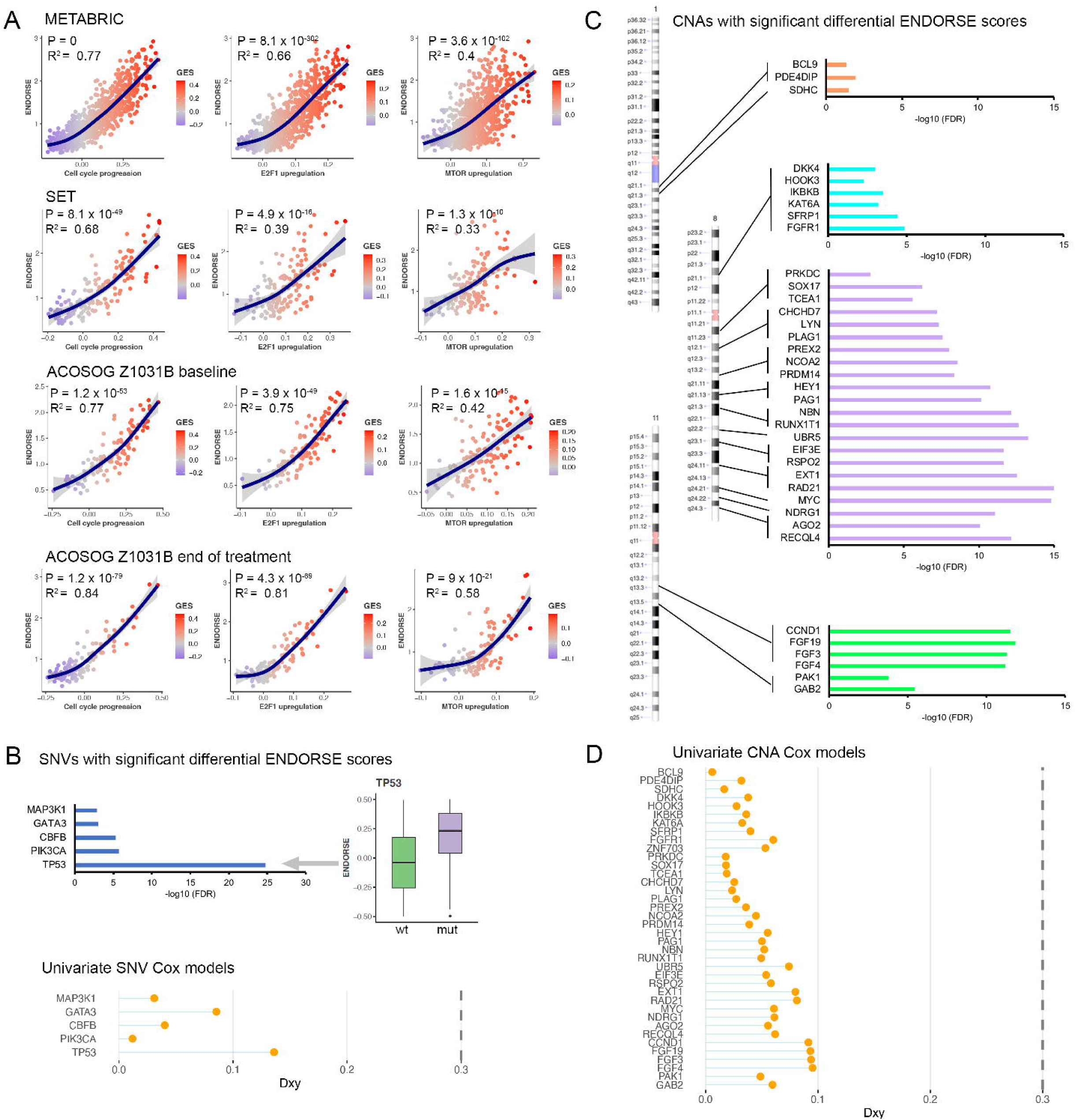
Biology of the high-risk tumors. A. Scatter plots displaying gene set enrichment scores (GES) of key pathways (X-axis) and ENDORSE scores (Y-axis). The cell cycle progression panel represents the hallmark G2M checkpoint signature, the E2F1 upregulation panel represents E2F1_UP.V1_UP oncogenic (C6) signature and the MTOR upregulation panel represents MTOR_UP.V1_UP oncogenic (C6) signature. Blue lines with shading indicate generalized additive model fits with 95% C.I., with R^2^ and p-values of the significant of the fit annotated on the panels. B. Barplots showing p-values from the ANOVA analysis of ENDORSE scores with mutation status as the grouping variable. The boxplot on the right shows difference in the ENDORSE scores between *TP53* mutant and wildtype tumors. The lollipop plot below shows Somer’s D_xy_ of the univariate Cox models for the SNVs, with the vertical dotted line indicating D_xy_ of the ENDORSE model. C. Ideograms showing mapped regions with copy number gains that are significant in ANOVA analysis of ENDORSE scores with copy number gain status as the grouping variables. Barplots on the right show p-values from the ANOVA analysis. D. Lollipop plot showing Somer’s D_xy_ of the univariate Cox models for the copy number gains, with the vertical dotted line indicating D_xy_ of the ENDORSE model.

We also analyzed the association between gene-level somatic mutations, including non-synonymous single-nucleotide variants (SNV) and copy number alterations, with the ENDORSE scores of the METABRIC ER+ tumors. We found a statistically significant association (FDR < 0.05) between the ENDORSE scores and SNVs of only five genes (Figure 6b, Supplementary Table 11). While *PIK3CA* mutations were found in ∼50% of all tumors, we found that ENDORSE scores were not significantly higher in tumors with non-synonymous *PIK3CA* variants or activating *PIK3CA* variants that guide the use of PI3K inhibitors (Supplementary Figure 3). Of the five significant genes, only tumors with *TP53* mutations showed a significantly higher ENDORSE score (Figure 6b). We then performed bootstrap analyses with the univariate Cox models of the significant genes and found that none of the SNV Cox models performed better than the ENDORSE model (Figure 6b).

Several gene-level amplifications were also associated with significant differences in ENDORSE scores (Figure 6c, Supplementary Figure 4). Interestingly, the significant amplifications were localized at chromosome 1q, 8p, 8q, or 11q, suggesting different genetic alterations affecting a recurring set of loci may be correlated with the emergence of resistance in the high-risk tumors (Figure 6c, Supplementary Table 12). Like the univariate SNV models above, the univariate copy number alteration models also performed poorly when compared to the ENDORSE model in bootstrap resampling analyses (Figure 6d).

## DISCUSSION

The criteria for classifying tumor as ER+ is based on a broad criteria of positive immunohistochemical staining of 1-100% of cell nuclei for the estrogen receptor^14, 32^. However, ER+ tumors are heterogeneous, both in terms of dependence on estrogen signaling for growth and survival and intrinsic or acquired resistance to endocrine therapy ^33, 34^. Therefore, optimal clinical management of each ER+ breast cancer depends on accurate prediction of response to endocrine therapy and selection of companions for endocrine therapy. Several genomic tests are available for classifying breast cancers into molecular subtypes ^35^ or assessing the likelihood of benefit from chemotherapy in early-stage, node-negative ER+ breast cancers ^7,30^. Results from the MINDACT and TAILORx studies^7, 8^ show that it is possible for node-negative, early-stage breast cancers to safely waive additional chemotherapy if they are predicted to be at a low risk of recurrence based on genomic signatures. However, these tests have not proven to be useful in the advanced and metastatic ER+ breast cancer setting. The default primary treatment for advanced ER+ breast cancer remains endocrine therapy, despite proven benefit from add-on targeted therapy or potential switch to chemotherapy. Therefore, the key challenge in advanced ER+ breast cancer is to stratify patients that will likely benefit from continued endocrine therapy and patients that are likely resistant to single-agent endocrine therapy and will benefit from selecting a different treatment strategy^36^.

To address this challenge, we have developed a new prognostic model to predict endocrine response in advanced ER+ breast cancers. We developed our model using invasive tumors from the METABRIC study that were ER+ and included node-positive, high-grade tumors. Our model addressed several challenges associated with the development of genomic biomarkers. Since the number of available features to train the genomic models tend to be much larger than the number of available samples (p >> n), it is quite easy to create complex prediction models that contain a large number of predictor variables. Often, such models perform very well in the training datasets, but the performance cannot be replicated in independent test datasets due to overfitting. A number of approaches have been proposed to address this issue. Broadly, these can be classified into unsupervised and supervised approaches. The unsupervised approach typically relies on grouping or clustering the samples into based on similarity of gene expression profiles, followed by analysis of association with survival outcomes^37^. Alternatively, a supervised approach is to perform dimensionality reduction prior to modelling the survival outcome or drug response using univariate or multivariate models^38^. Our model utilized the later strategy by using a regularized Cox model for feature selection, effectively reducing the dimensionality of the gene expression data. We further collapsed the genes into a signature and parameterized the final Cox model on the GES of the signatures. The rank-based approach to calculate GES also helped mitigate issues associated with batch effects and differences in methods for transcriptome profiling. We performed extensive performance evaluation of our model against other published signatures and clinical factors. Consistently, we found that the ENDORSE model was a better predictor than all other models in the training dataset (Figure 2). Moreover, ENDORSE clearly outperformed all other published signatures when they were applied to external validation datasets (Figures 3-5). Our results show that ENDORSE is a highly accurate and reproducible model that outperforms current approaches to predict endocrine response in metastatic ER+ breast cancer.

We also explored the biology of the ER+ tumors to identify possible mechanisms that are commonly shared by high-risk tumor. We found that high-risk tumors showed a consistent enrichment of pathways associated cell cycle progression and gain of PI3K/MTOR signaling pathways (Figure 6a). In addition, we observed consistent gain of the E2F1 signature, which may be associated with metastatic progression of breast cancers^39, 40^. We also observed loss of Rb1 activity, which has been associated with therapeutic resistance in ER+ breast cancers ^41, 42^

In addition to common pathway phenotypes shared across high-risk tumors, mutations in the *TP53* tumor suppressor genes were also significant (Figure 6b). Loss of function *TP53* variants have long been associated with aggressiveness and chemotherapeutic resistance in hormone-receptor negative breast cancers ^43, 44^. However, recent studies show that even though *TP53* mutations are infrequent in ER+ breast cancers, they have similar negative impact on patient outcome as hormone-receptor negative breast cancers ^45^. We also found recurrent copy number gains at chromosomes 8 and 11 to be associated with high-risk tumors (Figure 6c). Amplifications at these loci have been previously associated with aggressive and drug resistant cancers, and included several oncogenes such as *MYC*, *CCND1* and multiple fibroblast growth factors ^46, 47^. The survival models based on genomic alterations were clearly outperformed by ENDORSE; however, the recurrent nature of these alterations in high-risk tumors suggests further studies to investigate their role in promoting endocrine resistance are warranted.

Drugs that target CDK4/6 to inhibit cell cycle activation^48^, PI3K-inhibitors that target tumor with activating *PIK3CA* mutations^12^ and mTOR-inhibitors that prevent the activation of mTOR signaling and cell proliferation^13^ have been studied and approved for the treatment of advanced ER+ breast cancers in combination with endocrine therapy. However, patients must first advance on primary endocrine therapy, with or without additional CDK4/6 inhibitors, before they can be stratified in a different treatment arm. Therefore, identifying high-risk tumors with the ENDORSE model prior to first-line administration of single-agent endocrine therapy could help identify which cancers may be better suited for an add-on regimen or switching to chemotherapy. Thus, future clinical trials applying ENDORSE model may benefit from early and accurate prediction of endocrine response in advanced, metastatic ER+ breast cancers. This could ultimately help prolong survival of patients by stratifying in more appropriate treatment group.

## METHODS

### Data retrieval and pre-processing

METABRIC gene expression, phenotypic and survival data were retrieved using cBioPortal for cancer genomics^49^. Independent validation datasets used in this study were retrieved from the NCBI Gene Expression Omnibus database with the following accession IDs: SET ER/PR GSE124647^18^, TransCONFIRM GSE76040^19^, and ACOSOG Z1031B GSE87411^31^. For each gene expression dataset (log2 transformed), we removed genes with zero variance and summarized genes with multiple probes by mean expression and scaling of the expression levels of each gene to a mean of zero and standard deviation of one.

### Inclusion criteria for METABRIC training cohort

The METABRIC cohort contained a total of 2509 samples. Samples that met all of the following criteria were included in the training cohort: patients that were ER-positive and HER2-negative based on immunohistochemistry, patients that received hormone therapy but did not receive additional chemotherapy, patients that were either alive or died due to the disease and no other causes, and patients with complete survival and transcriptomic data. After filtering, 833 samples were retained for model construction.

### Empirical signature and ENDORSE model construction

The empirical gene signature was developed using a LASSO-regularized Cox proportional hazards models, with OS as the outcome variable^50^. The hazard function in the Cox model is defined as:

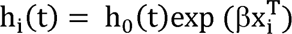

Where, X is a set of predictive gene expression features and h_0_ is an arbitrary baseline hazard function. The coefficient (β) for each predictor in the model can be estimated by maximizing the partial likelihood function L(β), defined as:

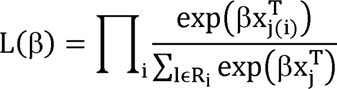

Where R_i_ is the set of indices of observations failing (events) at time t_i_. In the LASSO Cox model, the regularized coefficient is obtained by adding a penalty parameter λ to the log of the likelihood function.

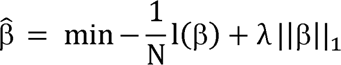

Where, l(β) = log L(β). The λ penalty parameter was determined using 10-fold cross-validation implemented in R package glmnet ^36, 37^. The optimal λ minimized model deviance.

We applied the model in a repeated (50 x 10-fold) cross validation framework. In each iteration, a set of ‘seed genes’ or features with positive coefficients in the regularized Cox model at a λ equal to one standard error from the minimum model deviance were identified. The seed genes were expanded to a redundant correlation network by adding all genes in the training transcriptome dataset that had Pearson’s correlation > 0.75 with any of the seed genes. Across all iterations, we identified the common set of features that were present in at least 50% of the correlation networks and defined this set of features as the empirical signature.

The ENDORSE model was defined as the hazard’s ratio of the Cox proportional hazards model fitted on OS data of the training cohort with two components: GES for the empirical gene signature and GES for the hallmark estrogen early response

signature.

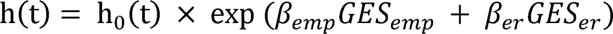

where, *emp* represents the empirical signature and *er* represents the estrogen response signature.

For each signature, the GES were calculated for individual samples using the GSVA package for R^53^ using the ssGSEA method^54^. The parameters for the ENDORSE model were obtained by fitting the model to the full training cohort of 833 samples, resulting in β_emp_ = 1.54 and β_er_ = -2.72.

### Models based on external signatures and clinical factors

Clinical features such as tumor grade and mutation count, along with scores from PAM50 and IntClust analyses were obtained directly from the METABRIC clinical annotations accompanying the transcriptome data and were directly utilized in univariate Cox models. Proliferation index based on the metaPCNA signature was calculated using the R-package ProliferativeIndex^27^.

We replicated the signatures and algorithms developed in the TransCONFIRM, SET ER/PR and 21-gene prognostic signature studies by following the methods described in the respective studies. The TransCONFIRM signature composed of 37 genes was implemented by performing hierarchical clustering of the gene expression data using these genes and cutting the tree (k=2) to stratify samples in high or low TransCONFIRM score categoties. The SET signature was implemented by calculating (the average expression of the 18-genes in the signature) – (the expression of 10 house-keeping genes) + 2. The 21-gene signature (ODX) score was calculated by following the unscaled risk score calculation reported by the study. *BAG1* transcript was missing from the METABRIC cohort and was not included in the unscaled score calculation. Since this transcript was uniformly missing on all samples, the relative risk scores could be compared across the samples.

### Cox model performance evaluation in training data

The predictive ability of ENDORSE and various other models were evaluated in the METABRIC training dataset using a bootstrap resampling analysis of the Cox regression models. The resampling was repeated 150 times for each model and a Somer’s D_xy_ rank correlation was calculated in each repeat. A final bias-corrected index of Somer’s D_xy_ was obtained as measure of the model’s predictive ability. The bootstrap resampling and calculations of the Somer’s D_xy_ were performed using the R package ‘rms’. Models based on SNVs and CNAs significantly associated with ENDORSE scores were also evaluated by obtaining Somer’s D_xy_ rank correlation metric of the univariate Cox model.

To compare each of the external signatures and clinical feature models with the ENDORSE model, we applied Vuong’s^55^ partial likelihood ratio test for non-nested Cox regression models calculated using the R package ‘nonnestcox’ (https://github.com/thomashielscher/nonnestcox/). The individual components of the ENDORSE model were compared to the full model using likelihood ratio tests for nested Cox models.

### Model validation in independent datasets

We compared the predictive performance of ENDORSE in multiple independent datasets. First, we integrated the training (METABRIC) and test (independent validation) datasets to perform batch correction using the ComBat function of the R package ‘sva ’^56^. Next, we calculated the GES for the ENDORSE signatures in the training (METABRIC) and test (independent validation) splits of the batch-corrected gene expression dataset. Then, the parameters of the ENDORSE Cox model were calculated on the batch-corrected training split with the OS information as the response variable. Finally, the parameterized Cox model was applied to the test split to obtain a predicted risk score.

In case of the SET ER/PR cohort, we used the predicted ENDORSE risk scores to stratify the patients into risk categories, with an ENDORSE score ≥ 2 representing the high-risk group, ≤1 representing the low-risk group and other intermediate values representing the medium risk group. We compared the significance of stratification of both OS and PFS curves based on ENDORSE, SET and TransCONFIRM scores using log-rank tests. Further, we compared the models using partial likelihood ratio tests for non-nested Cox models.

For the TransCONFIRM and ACOSOG cohorts, we compared the ENDORSE risk scores, SET and TransCONFIRM predictions with reported clinical variables, such as percentage of cells positive for Ki67 at the end of treatment and clinical outcomes using generalized linear models for continuous outcome variables or one-way ANOVA analysis for categorical outcomes.

### Biological features associated with ENDORSE scores

To determine the possible biological mechanisms associated with emergence of endocrine resistance and high ENDORSE risk scores, we evaluated the enrichment scores of various biological pathway and oncogenic signatures across the training and independent validation cohorts. We used the ssGSEA method to obtain GES for hallmark, curated (C2) and oncogenic signature (C6) gene sets from the molecular signatures database^57^. For each signature, we fitted a generalized additive model against the predicted ENDORSE score to obtain significance of the fit, R^2^ and proportion of variance explained by the model. None of the curated signatures were significant in the METABRIC analyses and were excluded from further consideration in the independent validation datasets.

Gene-level somatic SNV and CNV analyses were performed using data reported by the METABRIC study. SNV’s were retained based on a mutation frequency of ≥5 across all samples and limited to genes that are known cancer-related genes. Pathogenic *PIK3CA* variants associated with PI3K inhibitor sensitivity were obtained from the drug labels for alpelisib based on the SOLAR1 clinical trial^12^. Significant SNVs and CNVs were obtained using a one-way ANOVA analysis of the ENDORSE scores with mutation status as the factor.

### Data availability and code

All training and validation datasets used in this study are publicly available and listed under “data retrieval, preprocessing and analysis”. All analyses were performed in RStudio (1.2.5033, R 3.6.3). The sample code for reproducing the analyses in this study are available at https://osf.io/bd3m7/?view_only=da4f860bd2474745880944fce1d433b1

## Data Availability

All data used in this manuscript are publicly available and listed under "Methods" section of the manuscript

## ACKNOWLEDGEMENTS

Funding for this research was provided by the National Cancer Institute of the National Institutes of Health through the U54 grant 1U54CA209978.

## AUTHOR CONTRIBUTIONS

Conceptualization, Methodology, Investigation – AN, ALC, AHB; Data Curation, Formal Analysis, Software, Visualization, Validation – AN; Funding Acquisition, Resources, Supervision, Project Administration – ALC, AHB; Writing – original draft – AN; Writing – review & editing – AN, ALC, AHB.

