## Supplementary figures and images for "ENDORSE: a prognostic model for endocrine therapy response in advanced estrogen-receptor positive breast cancers"

### Supplementary Figure 1

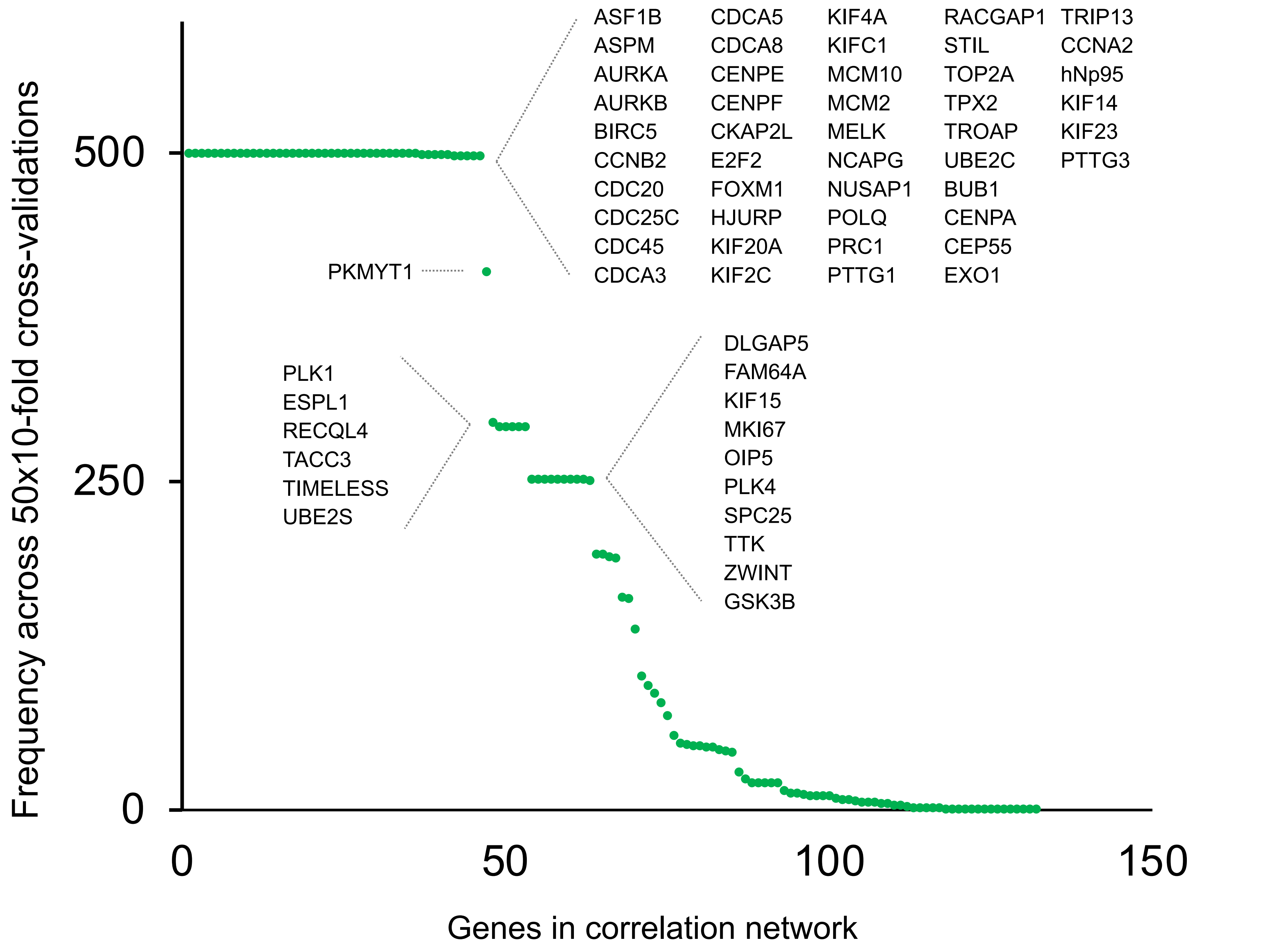

### Supplementary Figure 2

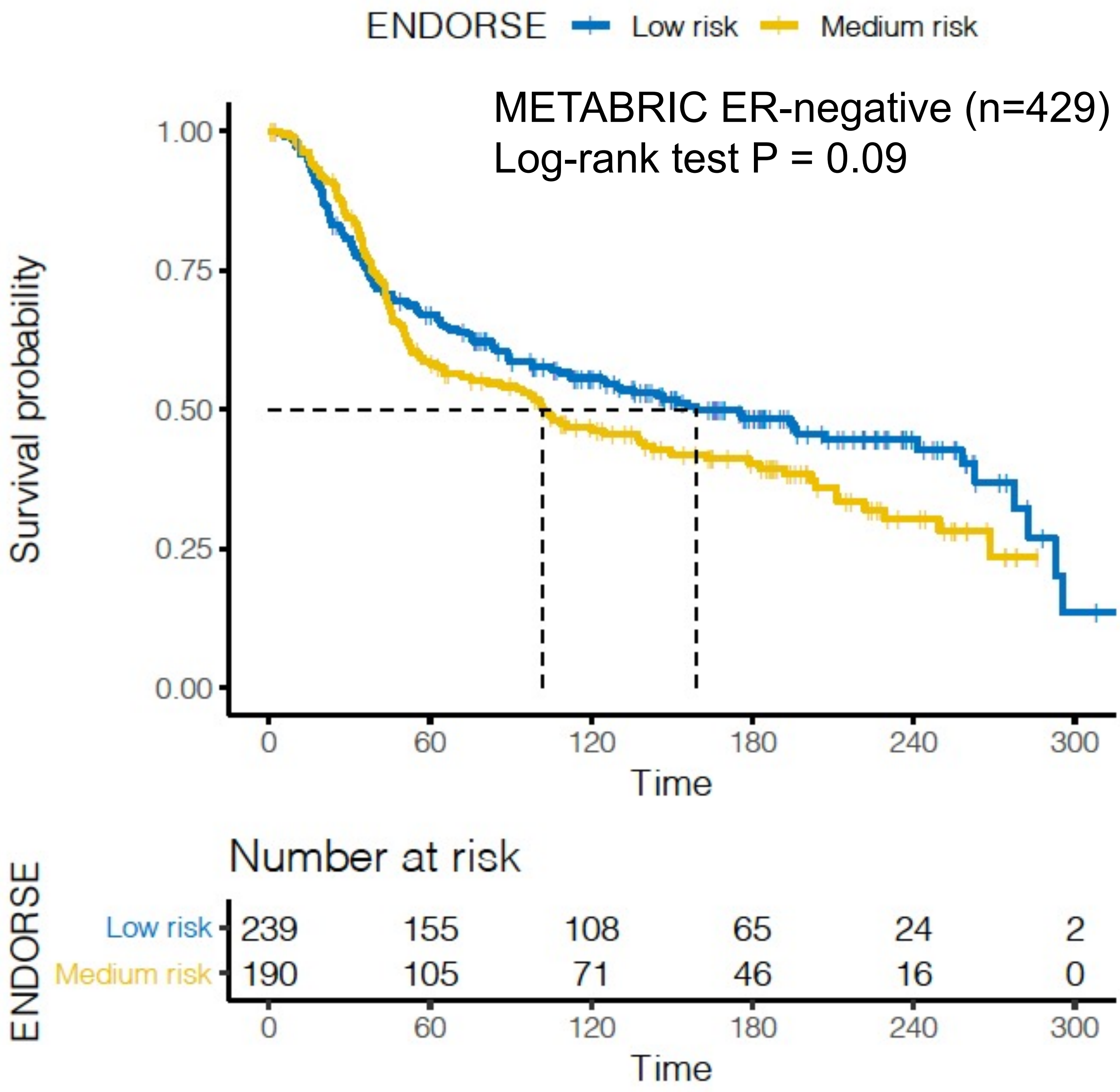

### Supplementary Figure 3

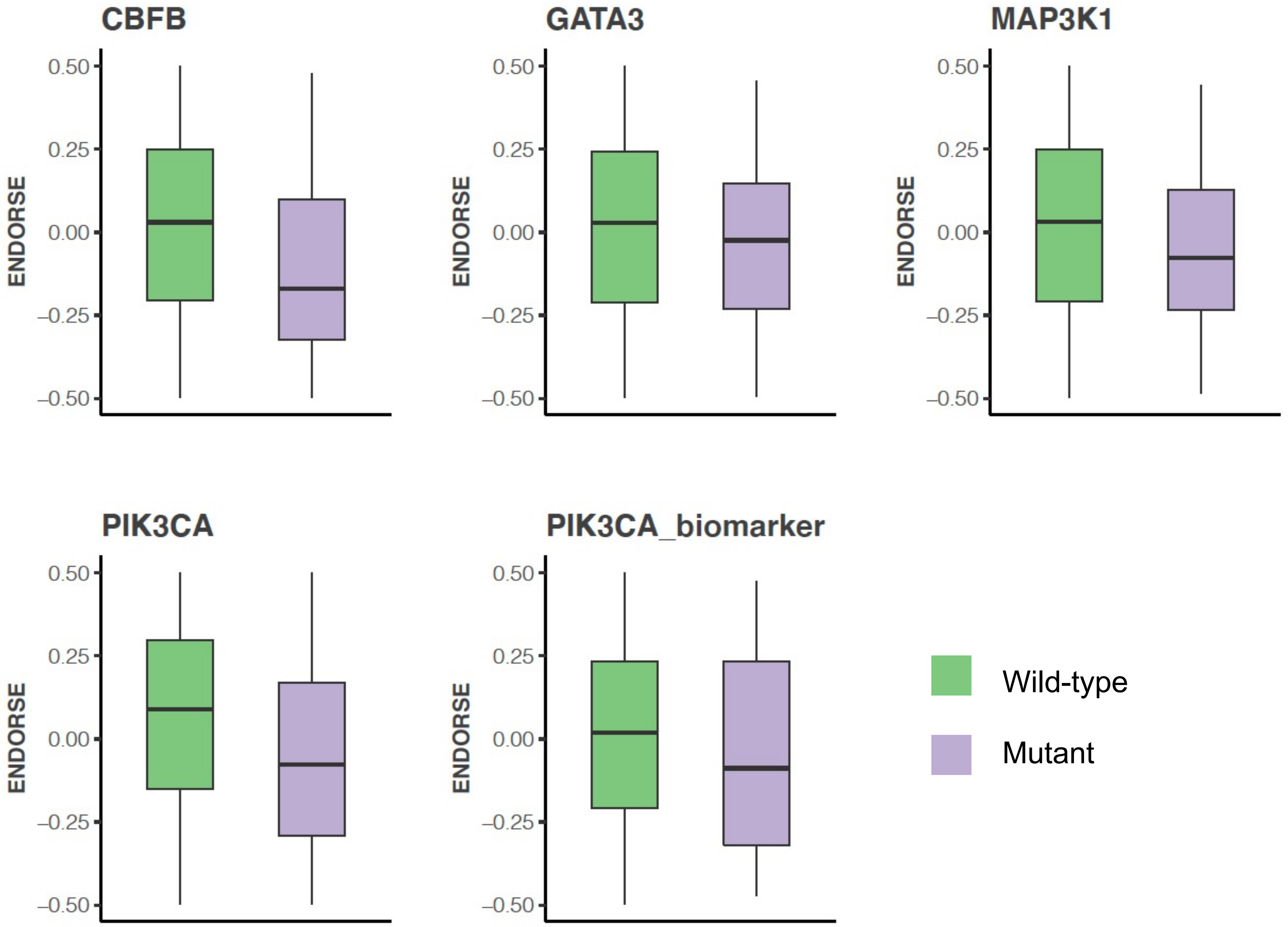

### Supplementary Figure 4

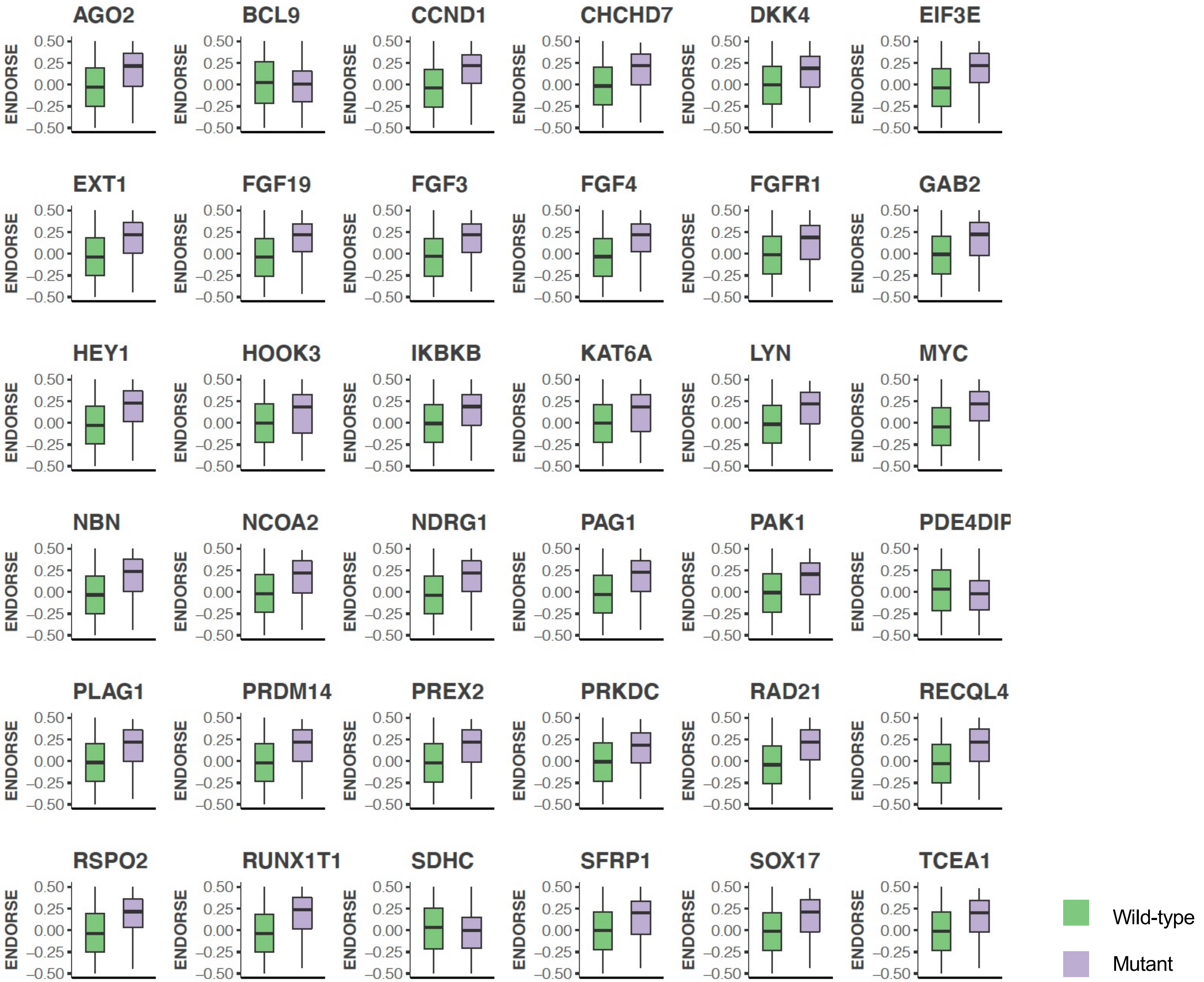
